# Understanding the comorbidities among psychiatric disorders, chronic low-back pain, and spinal degenerative disease using observational and genetically informed analyses

**DOI:** 10.1101/2025.02.28.25323099

**Authors:** Dan Qiu, Eleni Friligkou, Jun He, Brenda Cabrera-Mendoza, Mihaela Aslan, Mihir Gupta, Renato Polimanti

## Abstract

Psychiatric disorders and symptoms are associated with differences in pain perception and sensitivity. These differences can have important implications in treating spinal degenerative disease (SDD) and chronic low-back pain (CLBP). Leveraging data from the UK Biobank (UKB) and the All of Us Research Program (AoU), we investigated the effects linking psychiatric disorders (alcohol use disorder, anxiety, attention deficit hyperactivity disorder, bipolar disorder, cannabis use disorder, depression, opioid use disorder, posttraumatic stress disorder, and schizophrenia) to SDD and CLBP. We applied multi-nominal regression models, polygenic risk scoring (PRS), and one-sample Mendelian randomization (MR) to triangulate the effects underlying the associations observed. We also performed gene ontology and drug-repurposing analyses to dissect the biology shared among mental illnesses, SDD, and CLBP. Comparing individuals affected only by SDD (UKB N=37,745, AoU N=3,477), those affected only by CLBP (UKB N=15,496, AoU N=23,325), and those affected by both conditions (UKB N=11,463, AoU N= 13,451) to controls (UKB N=337,362, AoU N= 117,162), observational and genetically informed analyses highlighted that the strongest effects across the three case groups were observed for alcohol use disorder, anxiety, depression, and posttraumatic stress disorder. Additionally, schizophrenia and its PRS appeared to have an inverse relationship with CLBP, SDD, and their comorbidity. One-sample MR highlighted a potential direct effect of internalizing disorders on the outcomes investigated that was particularly strong on SDD. Our drug-repurposing analyses identified histone deacetylase inhibitors as targeting molecular pathways shared among psychiatric disorders, SDD, and CLBP. In conclusion, these findings support that the comorbidity among psychiatric disorders, SDD, and CLBP is due to the contribution of direct effects and shared biology linking these health outcomes. These pleiotropic mechanisms together with sociocultural factors play a key role in shaping the SDD-CLBP comorbidity patterns observed across the psychopathology spectrum.

## INTRODUCTION

Low back pain (LBP) encompasses a wide variety of types of pain with a cumulative point prevalence of 12% in the general adult population and a lifetime prevalence of approximately 40% globally.^1, 2^ Consequently, LBP is a leading cause of years lived with disability and productivity loss.^3^ Clinical management is driven by both the etiology and duration of LBP. The classification of LBP is broadly divided into radicular (also known as neuropathic), mechanical or nociplastic causes, or combinations of these etiologies.^2^ Chronic low back pain (CLBP), defined as LBP lasting more than 12 weeks duration, constitutes a rising public health burden particularly in older adults, with an estimated 12-month prevalence approaching 36% in those over age 60.^4^

In most cases of CLBP, identifying a specific cause has historically been challenging.^5^ Although the increasing adoption of advanced imaging and interventional modalities has improved diagnostic accuracy for structural causes of CLBP,^2^ several factors preclude successful treatment of CLBP. For example, the high prevalence of advanced spinal degenerative changes in asymptomatic individuals confounds the identification of specific pain-generating structures in those with multifocal disease.^6, 7^ Additionally, CLBP and other chronic pain conditions arise from a complex interplay between biological, psychological, and social factors, termed the biopsychosocial model of pain.^2, 8, 9^ Behavioral patterns, social determinants such as educational attainment, genetic factors, and psychiatric comorbidities have all been demonstrated to have significant and complex relationships with CLBP.^10, 11^

Psychiatric disorders in particular are associated with differences in pain perception,^12, 13^ but the mechanisms underlying these relationships remain unclear and there is divergent evidence regarding pain sensitivity in these populations.^12, 14^ Historically, psychiatric conditions were believed to be more prevalent among individuals without identifiable structural causes of CLBP.^15^ However, psychiatric comorbidities have recently been associated with advanced symptoms, poor outcomes, and increased resource utilization in patients being treated for CLBP due to specific degenerative disease conditions.^16–18^ Emerging evidence raises the hypothesis that the relationship between psychiatric comorbidities and CLBP may be mediated by shared developmental and neural mechanisms.^19^

CLBP associated with both spinal degenerative disease (SDD) and psychiatric comorbidities represents a unique and increasingly prevalent therapeutic challenge, because no framework currently exists for characterizing the relative contributions, relationships, and underlying mechanisms of psychiatric and structural factors. Understanding these dynamics has the potential to enable personalized and rational selection from among the myriad behavioral, pharmacologic, integrative, and surgical treatment approaches to CLBP.^2^ The present study conducted observational and genetically informed analyses to explore how psychiatric disorders and symptoms are differentially associated with CLBP and SDD in two independent cohorts comprising a total of 559,487 participants.

## METHODS

### Study design and participants

This study explored the associations between psychiatric disorders, SDD, and CLBP using data from the UK Biobank (UKB)^20^ and the All of Us Research Program (AoU)^21^ through both observational and genetically informed analyses (Figure 1). UKB received ethical approval from the National Health Service National Research Ethics Service Northwest (reference: 11/NW/0382). AoU was approved by its Institutional Review Board. UKB and AoU obtained informed consent from all participants enrolled. The study was performed in agreement with the Helsinki Declaration.

**Figure 1.**
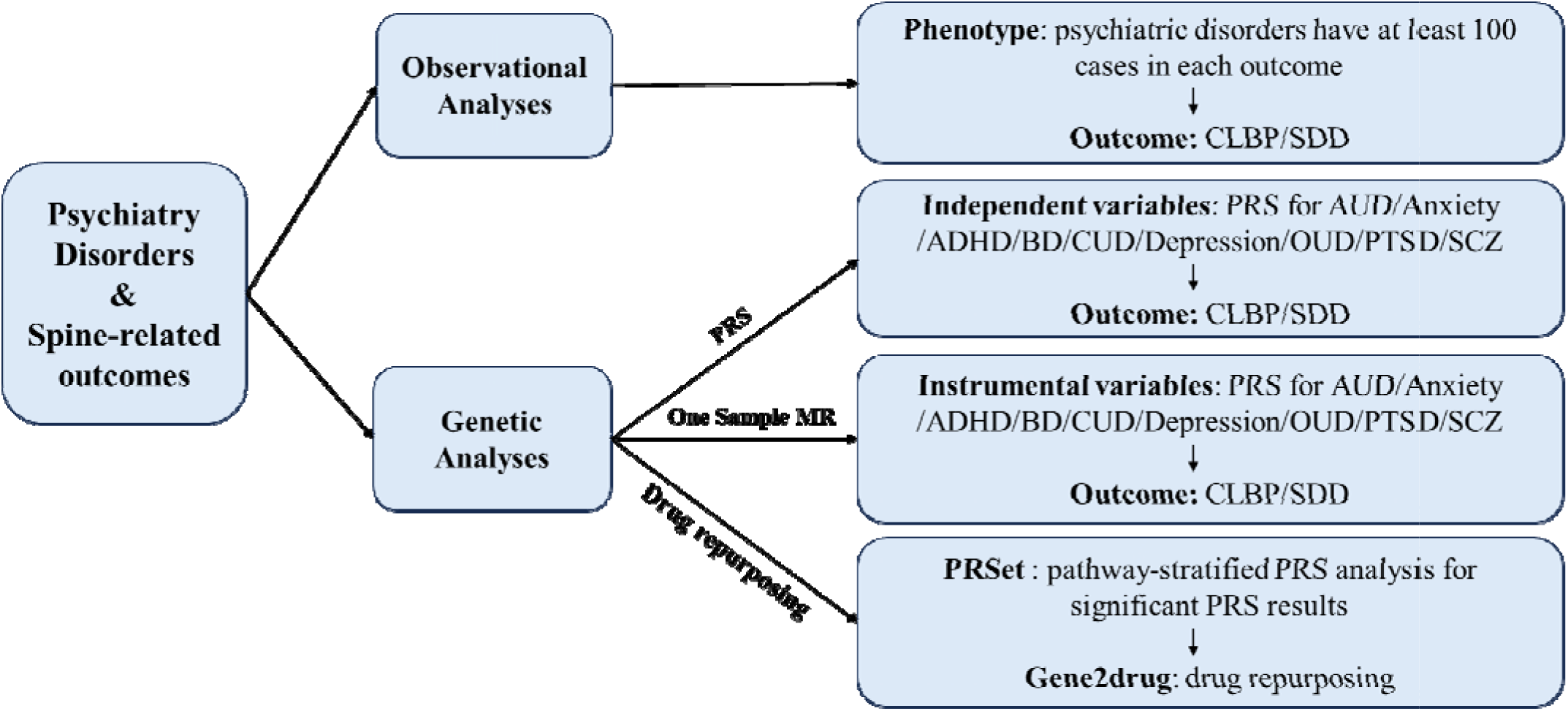
Study overview. *Note:* AUD: Alcohol Use Disorder; ADHD: Attention Deficit Hyperactivity Disorder; BD: Bipolar Disorder; CUD: Cannabis Use Disorder; OUD: Opioid Use Disorder; PTSD: Posttraumatic Stress Disorder; SCZ: Schizophrenia; CLBP: Chronic low back pain; SDD: Spinal degenerative disease; PRS: Polygenic risk score; MR: Mendelian Randomization.

### Phenotyping

The present study investigated the following psychiatric disorders: alcohol use disorder (AUD), anxiety, attention deficit hyperactivity disorder (ADHD), bipolar disorder (BD), cannabis use disorder (CUD), depression, opioid use disorder (OUD), posttraumatic stress disorder (PTSD), and schizophrenia (SCZ) were investigated in this study. In UKB, psychiatric disorders were assessed using “first occurrence” data, which combined information from primary care records, hospital admission records, death register records, and self-reported surveys.^20^ In AoU, these psychiatric disorders were identified from the “conditions” domain using electronic health record (EHR) data standardized by the Observational Medical Outcomes Partnership Common Data Model (OMOP CDM).^21^ To maximize the sample size available In both UKB and AoU, SCZ cases also included individuals reporting psychotic symptoms (e.g., visual hallucinations, auditory hallucinations, delusions of reference, and delusions of persecution) as similarly to what performed previously.^22^ Specific UKB field IDs and AoU concept IDs for the independent variables were detailed in Table S1.

To define CLBP and SDD outcomes, we used UKB EHRs available from primary and secondary diagnoses, medication records, and procedure records. In the AoU cohort, CLBP and SDD were identified from the “conditions” domain using EHR data standardized by the OMOP CDM.

For CLBP, participants were identified based on ICD-10 codes M54.3-M54.5, which pertain to dorsalgia. For SDD, identification was based on both diagnoses and operation records. Specifically, diagnoses included various ICD-10 codes such as spondylosis (M47), as well as ICD-9 codes including spinal stenosis (720), spondylosis and allied disorders (721), and several others related to disc disorders and spinal curvature. Operative procedures were coded according to the Office of Population Censuses and Surveys Classification of Interventions and Procedures, version 4 (OPCS4). Relevant procedures included primary microdiscectomy of the lumbar intervertebral disc (V33.7), primary posterior laminectomy decompression of the lumbar spine (V25.4), and various other specified and unspecified decompression and excision operations related to both lumbar and cervical spines. Injection procedures, such as therapeutic lumbar epidural injections (A52.1) and radiofrequency controlled thermal denervation of spinal facet joints (V48.5), were also included as part of the identification for SDD.

To define CLBP and SDD, we excluded participants affected by medical conditions and treatments that could present similar symptoms due to different pathophysiological processes (Table S1). For example, individuals with cancers such as lung and colon cancer were excluded. Additionally, those with infections like osteomyelitis or pyogenic arthritis, as well as neurological conditions such as multiple sclerosis, were not included. Participants taking medications like plaquenil and indomethacin, and individuals associated with surgical procedures affecting the spine (OPCS4 codes: A57.4, A57.8, V44.4) were also excluded.

The detailed definitions and specific codes used for CLBP and SDD phenotyping are provided in Table S1. Based on these definitions, participants were categorized into four groups: i) those with both CLBP and SDD (CLBP+SDD cases), ii) those with only CLBP (CLBP-only cases), iii) those with only SDD (SDD-only cases), and iv) those with neither condition (i.e., controls).

Information regarding sex, age, smoking status, and body mass index (BMI) were also extracted from UKB and AoU (Table S1).

### Genetic data

Genome-wide data from previous large-scale studies were used to investigate further the genetic effects linking psychiatric disorders to CLBP and SDD. To avoid sample overlap bias in the polygenic risk score (PRS) analyses, we investigated genome-wide association studies (GWAS) that did not include the UKB cohort and the AoU cohort. Because of the limited sample size available for other ancestry groups, we used genome-wide data generated from individuals of European descent. Specifically, schizophrenia,^23^ bipolar disorder,^24^ ADHD,^25^ and PTSD,^26^ genome-wide data were obtained from the Psychiatric Genomic Consortium (PGC). AUD, depression, CUD, and OUD GWAS were obtained from a meta-analysis including multiple cohorts such as PGC, Million Veteran Program (MVP), iPSYCH, and deCODE.^27–30^ Leveraging data generated from a previous study,^31^ our group also generated two anxiety GWAS meta-analyses: one excluding UKB cohort (generated from the meta-analysis of PGC, iPSYCH, MVP, FinnGen, and AoU samples) and another excluding AoU cohort (generated from the meta-analysis of PGC, iPSYCH, MVP, FinnGen, and UKB samples), ensuring no overlap in participant inclusion across the datasets.

These genome-wide association statistics were integrated with individual-level genetic data from UKB and AoU (Fig. S1). UKB genetic information was processed considering quality control (QC) measures and ancestry assignments as previously described by the Pan UKB initiative.^32, 33^ For the AoU dataset, whole-genome sequencing data were extracted using the ACAF threshold callset, with associated quality control criteria and ancestry inference described in previous literature.^34^ Additionally, we excluded variants with a call rate below 95%, a minor allele frequency (MAF) below 1%, and a Hardy-Weinberg equilibrium P-value less than 1×10^-6^ to ensure data integrity.

### Statistical analyses

To investigate the associations between psychiatric disorders, CLBP, and SDD, we conducted multinomial logistic regression analysis on phenotypic data from UKB and AoU. Specifically, we tested psychiatric disorders by comparing CLBP+SDD cases, CLBP-only cases, and SDD-only cases with respect to controls. To maximize UKB and AoU sample size, we considered psychiatric disorders with at least 100 cases in each of the CLBP and SDD categories (Table S2). The regression analyses were conducted considering two models: one including only age and sex as covariates and the other including age, sex, BMI, smoking status, and genetically inferred ancestries. Bonferroni multiple testing correction was applied to account for the number of tests performed (p<0.003). To estimate differences across psychiatric disorders and case groups, we applied a z-test to compare regression coefficients and their standard errors.

As mentioned above, genetic analyses were performed only in individuals of European descent because of the lack of large-scale samples informative of other population groups. Leveraging large-scale GWAS of psychiatric disorders, PRS were estimated using the Polygenic Risk Score–Continuous Shrinkage (PRS-CS) method,^35^ using the 1000 Genomes European-ancestry populations as the linkage disequilibrium (LD) reference panel.^36^ PRS-CS weights were then used to estimate individual PRS for UKB and AoU participants through the PLINK 1.9 --score command.^35^

The associations between psychiatric PRS and the outcomes were assessed using multinomial logistic regression, with the psychiatric PRS as the independent variable and CLBP and SDD as the outcome. Initially, we tested the nine psychiatric PRS into the model separately.

Subsequently, we conducted a multivariable analysis entering the nine psychiatric PRS into the same regression model. In these analyses, we used age, sex, and the top 10 within-ancestry principal components (PCs) as covariates. We also conducted further analyses, adding smoking status and BMI to the initial set of covariates.

To further investigate the associations among psychiatric disorders, CLBP, and SDD, we performed one-sample MR.^37^ Using psychiatric PRS as instrumental variables, we performed an MR analysis in both UKB and AoU European-ancestry participants applying the two-stage least-squares (2SLS) estimator available in ivreg package in R.^38^ Sex, age, smoking status, BMI and the top 10 within-ancestry PCs were included as covariates. Because the outcome in the models was a multi-classified variable (CLBP+SDD, CLBP-only, SDD-only, and controls), we created three binary variables corresponding to each case definition and conducted logistic regression in the second stage. Bonferroni multiple testing correction was applied to account for the number of MR tests performed (p<0.002). To ensure the reliability of our findings, we also conducted weak-instrument and Wu-Hausman tests to examine whether the instrumental PRS was weakly correlated with the exposure variables.^37^

To investigate the biological mechanisms underlying the association of psychiatric PRS with CLBP and SDD, PRSet enrichment analysis was performed testing 10,542 Gene Ontology (GO) terms available from the Molecular Signatures Database (MSigDB) using PRSice-2.^39, 40^ PRSet analysis was conducted using the full genome-wide information clumped considering a 1000 kb window and LD r^2^=0.1. Bonferroni correction was applied to define significant GOs after multiple testing correction (p<4.74Ξ10^-6^).

Then, a drug repurposing analysis was performed using Gene2drug approach.^41^ This permitted us to translate Bonferroni-significant GO enrichments into potential molecular targets available from the Connectivity Map.^42^ To decrease the redundancy among GO terms before entering them into the Gene2drug analysis, we used the REVIGO algorithm^43^ considering a uniqueness score greater than 0.7, UniProt as the reference database, and the SimRel method for semantic similarity measurement.

## RESULTS

### Observational analyses

A total of 402,072 UKB participants and 157,415 AoU participants were available for the observational analyses (Table 1). Considering psychiatric disorders with more than 100 cases in each cohort (Table S2), we investigated CLBP and SDD with respect to AUD, anxiety, depression, PTSD, and SCZ in UKB and AUD, anxiety, BD, depression, and PTSD in AoU.

**Table 1.**
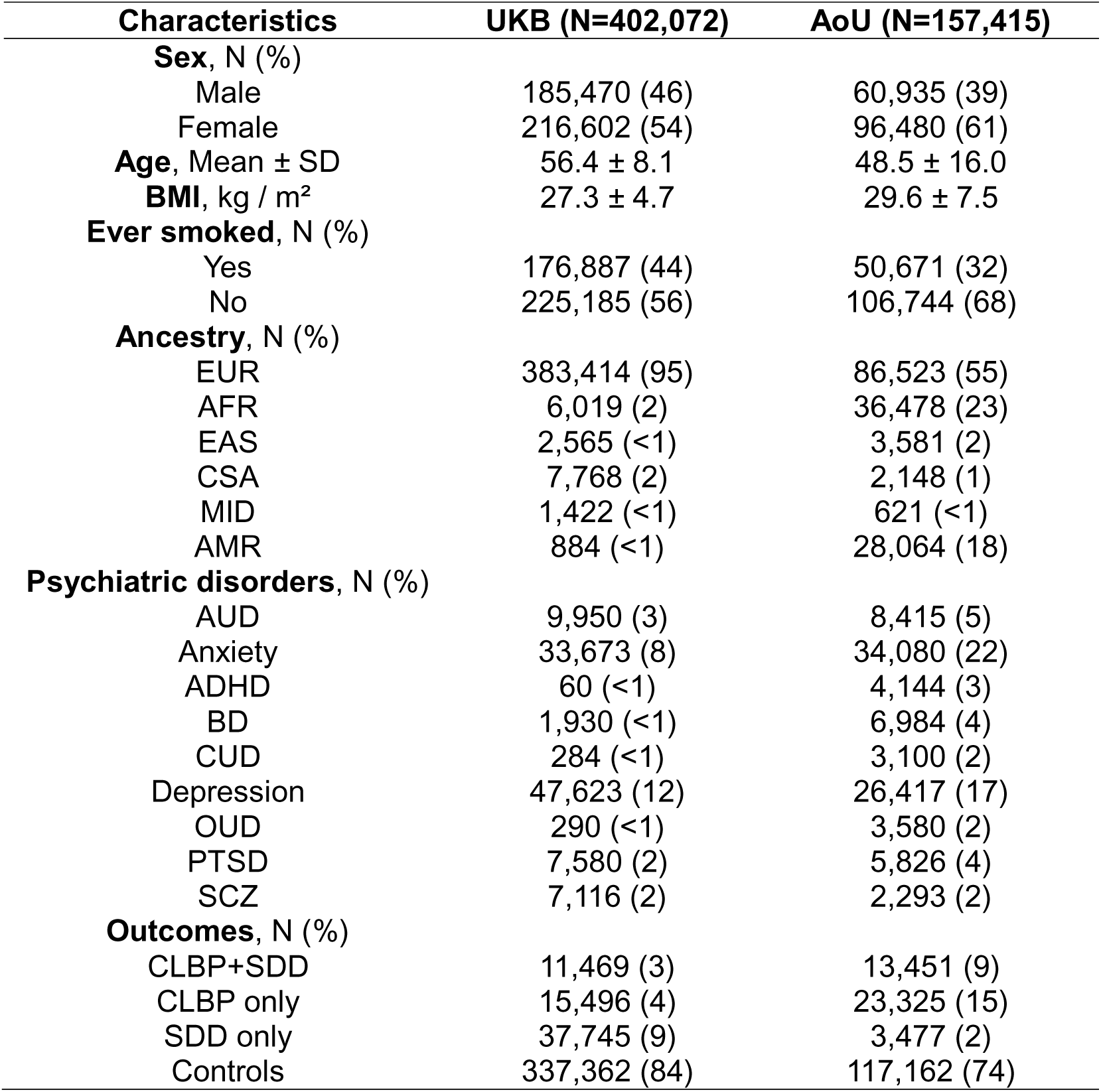
Characteristics of participants available from the UK Biobank and the All of Us Research Program.

In UKB, anxiety, AUD, depression, and PTSD were associated with increased CLBP+SDD, CLBP-only, and SDD-only case status after Bonferroni correction in both the age-sex model and in the fully adjusted model (p<0.003; Figure 2, Table S3). Comparing effect sizes in the fully adjusted model, we observed Bonferroni significant differences among psychiatric disorders (difference-p<0.002; Table S4). Because SCZ showed an inverse relationship with CLBP/SDD case status (Figure 2, Table S3), its effect size was statistically different from those observed for the other four psychiatric disorders investigated in UKB (Table S4). Quantitative differences were also observed among these psychiatric disorders (Table S4), especially because of the strong associations of depression with CLBP+SDD (β=0.720, 95%CI=0.670-0.769) and SDD-only (β=0.461, 95%CI=0.430-0.493) and between PTSD and CLBP-only (β=0.790, 95%CI=0.703-0.877). These two psychiatric disorders also showed differences among CLBP/SDD case status (Figure 2; Table S4). In PTSD, CLBP-only association was statistically stronger than the ones with CLBP+SDD (difference-p=1.77Ξ10^-4^) and SDD-only (difference-p=1.24 Ξ10^-24^). Conversely, for depression, CLBP+SDD association was statistically stronger than CLBP-only (difference-p=1.08Ξ10^-25^) and SDD-only (difference-p=4.22Ξ10^-18^). Anxiety showed an association pattern across CLBP/SDD case status similar to depression (Figure 2, Table S4).

**Fig. 2.**
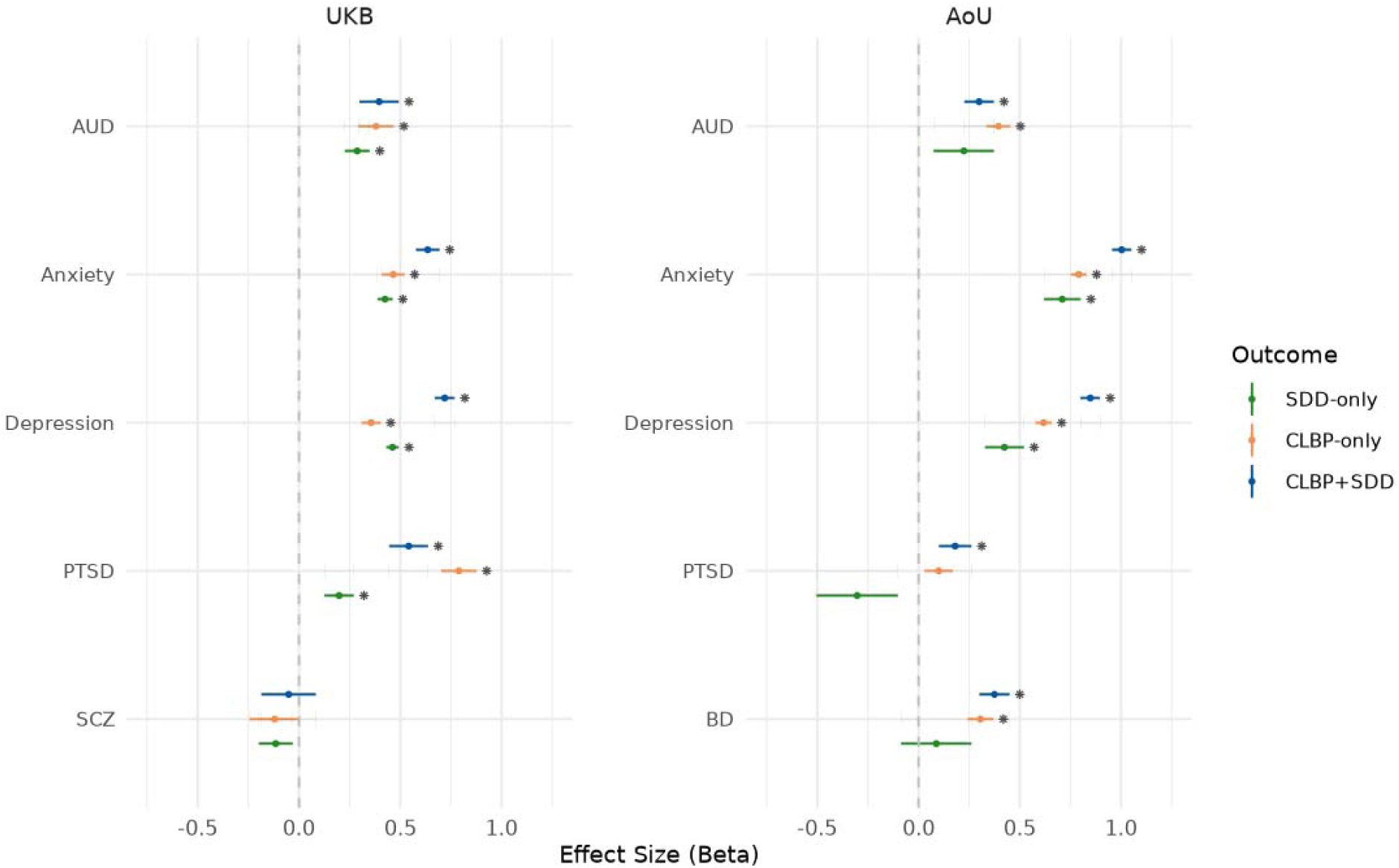
Association of psychiatric diagnoses (AUD: Alcohol Use Disorder; ADHD: Attention Deficit Hyperactivity Disorder; BD: Bipolar Disorder; CUD: Cannabis Use Disorder; OUD: Opioid Use Disorder; PTSD: Posttraumatic Stress Disorder; SCZ: Schizophrenia) with the comorbidity combinations between chronic low back pain (CLBP) and spinal degenerative disease (SDD) in UK Biobank (left panel) and All of Us Research Program (right panel). Bonferroni significant associations (p<0.003) are indicated with an asterisk. Full results are available in Table S3.

In AoU, no differences were observed between age-sex model and the fully adjusted model (Fig 2, Table S3). In both, AUD, anxiety, and depression were associated with increased status of the three CLBP/SDD case definitions after Bonferroni correction (p<0.003). BD was Bonferroni-significantly associated with increased CLBP+SDD and CLBP-only, but not with SDD-only (p>0.05). PTSD showed a Bonferroni significant association with increased CLBP+SDD (β=0.180, 95%CI=0.1-0.261), nominally significant associations with increased CLBP-only (β=0.099, 95%CI=0.03-0.169) and reduced SDD-only (β=-0.303, 95%CI=-0.504 – -0.102). Comparing effect sizes in the fully adjusted model, we observed Bonferroni significant differences among psychiatric disorders and across CLBP/SDD case definitions (Table S4).

Comparing UKB and AoU results, the main difference was observed with respect to PTSD associations with CLBP/SDD case definitions where UKB associations were much stronger than AoU. Additionally, UKB-PTSD was associated with increased CLBP/SDD case definitions, while AoU-PTSD was inversely related to SDD-only (Figure 2). In both UKB and AoU, anxiety and depression showed the strongest association with CLBP+SDD comorbidity, while small differences were present in AUD association strength among CLBP/SDD case definitions (Figure 2).

We also conducted ancestry-stratified analysis in both UKB and AoU samples. While most ancestry-specific associations were consistent with the ones observed in the cross-ancestry analyses (Table S5), we observed ancestry differences (Table S6) with the strongest ones related to AUD relationship with CLBP/SDD case definitions in UKB. Applying Bonferroni multiple testing correction (p<5.56×10^-4^) to the results of the fully adjusted model, AUD was inversely associated with CLBP-only (β =-3.183) and SDD-only (β=-2.925) in UKB participants of East Asian descent and with CLBP-only (β =-2.717) in UKB participants of European descent. Conversely, EUR associations of AUD with CLBP-only (β=0.386) and SDD-only (β=0.290) were consistent with the ones observed in the cross-ancestry analysis (CLBP-only β=0.380; SDD-only (β=0.287).

### Polygenic risk scoring

In UKB (325,975 unrelated participants of European descent), AUD, anxiety, ADHD, CUD, depression, OUD, and PTSD PRS showed Bonferroni significant associations (p<0.002) with increased CLBP/SDD case definitions (i.e., CLBP+SDD, CLBP-only, and SDD-only) in the model accounting for age, sex, and within-ancestry PCs and the model also including smoking and BMI (Figure 3, Table S7). Conversely, BD PRS was associated only with SDD-only status with an effect size (β=0.056, 95%CI=0.024-0.088) smaller than those observed for the other PRS mentioned. SCZ PRS also showed small effect sizes, but it also presented different effect directions. Specifically, SCZ PRS was associated with decreased CLBP+SDD (β=-0.006, 95%=-0.009 – -0.003) and increased CLBP only (β=0.012, 95%CI=0.009-0.016). To control for the shared genetic effects among the psychiatric disorders investigated, we conducted a multivariable analysis entering the PRS in the same model. In the fully adjusted multivariable model, only ADHD and depression PRS showed Bonferroni significant associations (p<0.002) with increased CLBP+SDD, CLBP, and SDD-only (Figure 3, Table S8). Conversely, AUD PRS was related only to increased CLBP+SDD (β=0.221, 95%CI=0.131-0.311) and CLBP (β=0.134, 95%CI=0.056-0.213). Anxiety and PTSD PRS were associated with increased SDD-only (β=0.134, 95%CI=0.071-0.197) and CLBP+SDD (β=0.187, 95%CI=0.080-0.294), respectively.

**Fig. 3.**
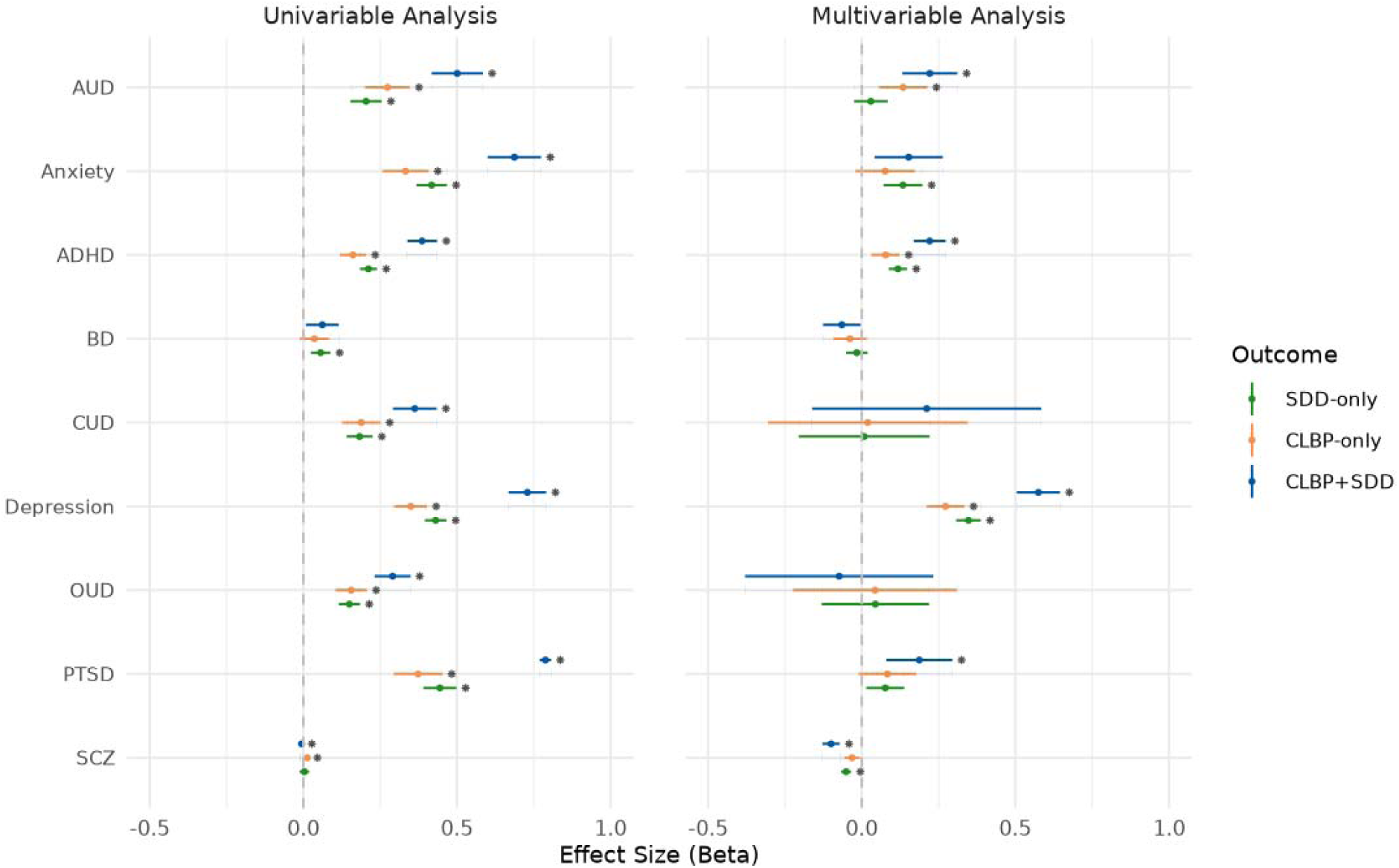
Associations of psychiatric polygenic risk scores (PRS; AUD: Alcohol Use Disorder; ADHD: Attention Deficit Hyperactivity Disorder; BD: Bipolar Disorder; CUD: Cannabis Use Disorder; OUD: Opioid Use Disorder; PTSD: Posttraumatic Stress Disorder; SCZ: Schizophrenia) with the comorbidity combinations between chronic low back pain (CLBP) and spinal degenerative disease (SDD) in UK Biobank. Single PRS associations are reported on the left panel, while the results of the multivariable PRS analysis are reported on the right panel. Bonferroni significant associations (p<0.002) are indicated with an asterisk. Full results are available in Tables S8 and S9.

Interestingly, accounting for the polygenic risk of other psychiatric disorders uncovered a Bonferroni-significant inverse relationship of SCZ PRS with both CLBP+SDD (β=-0.100, 95%=-0.128 – -0.072) and SDD-only (β=-0.051, 95%CI=-0.068 – -0.035). Comparing effect sizes obtained among the Bonferroni-significant PRS associations observed in the multivariable analysis, the strongest differences were mostly related to the strength of the association between depression PRS and CLBP+SDD (β=0.575, 95%CI=0.505-0.645), which was larger than the ones observed for CLBP and SDD-only and across the other psychiatric disorders (Table S9).

Because of the limited sample size available in AoU (48,994 unrelated AoU participants of European descent), we observed a reduced number of Bonferroni significant PRS associations (Table S7) compared to UKB. Specifically, considering the fully adjusted model, anxiety, ADHD, depression, and PTSD PRS showed Bonferroni significant associations with increased CLBP+SDD (p<0.002; Table S7). In the multivariable PRS analysis, we observed two Bonferroni significant PRS associations (Table S8). Depression PRS was associated with increased CLBP+SDD (β=0.275; 95%CI=0.100-0.450), while this relationship was inverse for SCZ PRS (β=-0.134; 95%CI=-0.211 – -0.058).

### One sample Mendelian randomization

In UKB, our one-sample MR analysis highlighted Bonferroni-significant causal effects (p<0.002) of AUD, anxiety, ADHD, CUD, depression, OUD, and PTSD on the three CLBP/SDD case definitions (Figure 4, Table S10). Generally, within each psychiatric disorder, the strongest effects were observed with respect to SDD-only. This difference was particularly strong for anxiety (SDD-only β =0.032, 95%CI=0.028-0.036), depression (SDD-only β=0.032, 95%CI=0.030-0.035), and PTSD (SDD-only β=0.034, 95%CI=0.029-0.038). In line with this specific relationship, BD effect – although small – was only significant with respect to SDD-only (β=0.004, 95%CI=0.002-0.007).

**Fig. 4.**
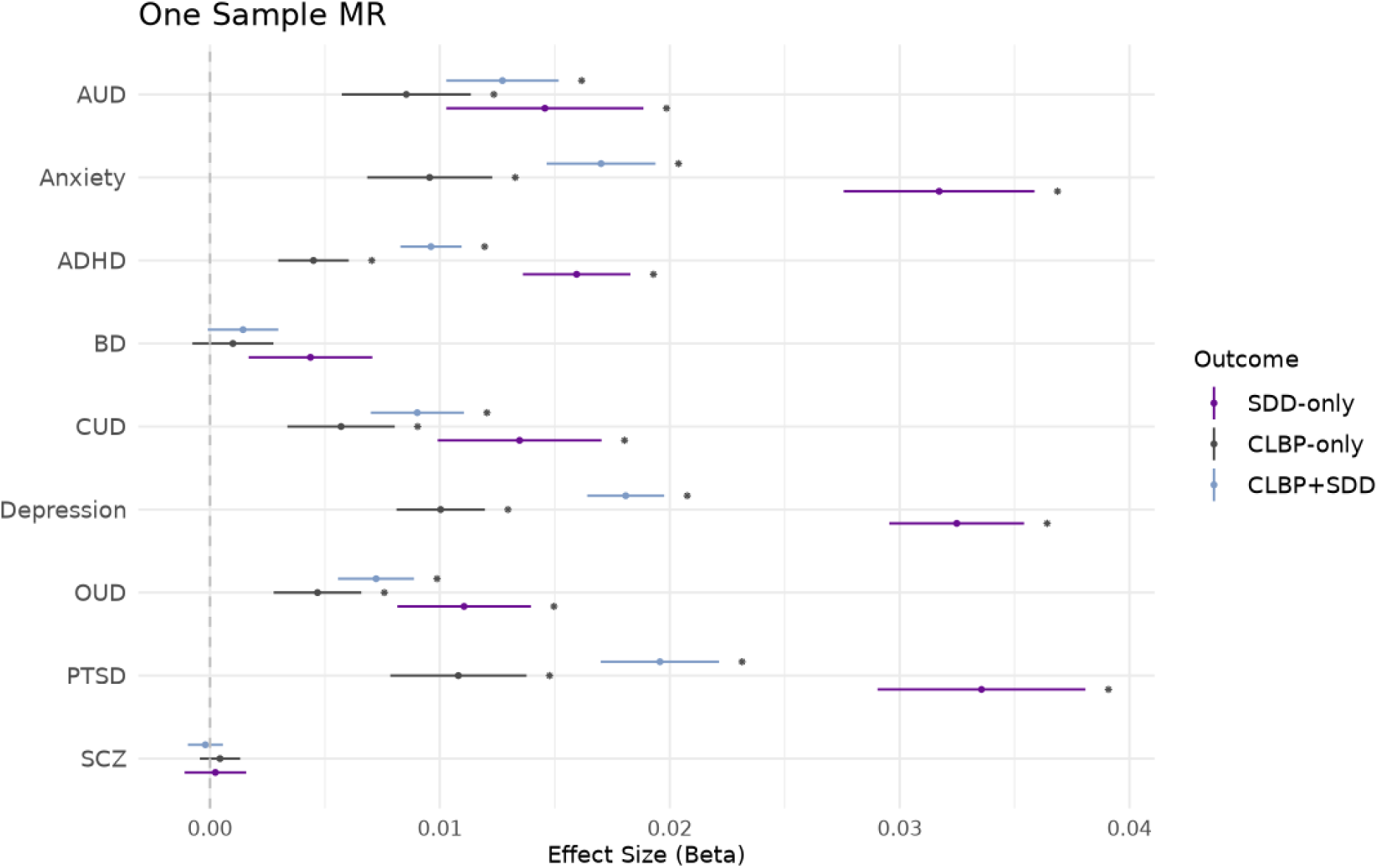
One-sample Mendelian randomization linking psychiatric disorders (AUD: Alcohol Use Disorder; ADHD: Attention Deficit Hyperactivity Disorder; BD: Bipolar Disorder; CUD: Cannabis Use Disorder; OUD: Opioid Use Disorder; PTSD: Posttraumatic Stress Disorder; SCZ: Schizophrenia) to comorbidity patterns between chronic low back pain (CLBP) and spinal degenerative disease (SDD) in UK Biobank. Bonferroni significant associations (p<0.002) are indicated with an asterisk. Full results are available in Table S10.

Because of the limited AoU sample size, we observed a Bonferroni-significant effect (p<0.002) only from depression to CLBP+SDD (β=0.219, 95%CI=0.109-0.329). This outcome also showed nominally significant relationships (p<0.05) with anxiety (β =0.192, 95%CI=0.065-0.319), PTSD (β=0.307, 95%CI=0.094-0.519) and ADHD (β=0.990, 95%CI=0.170-1.810). Depression also had a nominally significant effect on CLBP-only (β=0.165, 95%CI=0.031-0.299). Differently from UKB, psychiatric disorders appeared to affect more strongly CLBP+SDD than the other outcomes investigated in AoU (Table S10).

In both UKB and AoU, the one-sample MR sensitivity analyses of the Bonferroni-significant results confirmed the lack of weak instrument bias and the informativeness of the models, demonstrating the robustness of the effects observed.

### Gene ontology and drug repurposing analyses

To uncover biological processes, molecular functions, and cellular components underlying the PRS associations observed, we conducted a pathway-based analysis using PRSet approach.^40^ Considering Bonferroni correction accounting for the number of tests performed (N=10,542, p<4.74×10^-6^), we identified many significant GO terms (Table S11) underlying the association of CLBP/SDD case definitions with PRS related to anxiety (UKB N=227), AUD (UKB N=311), depression (UKB N=421, AoU N=201), PTSD (UKB N=13), and SCZ (UKB N=67, AoU N=4).

Considering Bonferroni significant GOs in both UKB and AoU, depression PRS associations with CLBP/SDD case definitions were enriched for “chemical homeostasis” (UKB p<1×10^-300^, AoU p=2.96×10^-7^), “generation of neurons” (UKB p<1×10^-300^, AoU p=1.17×10^-13^), “developmental growth” (UKB p=1.28×10^-233^, AoU p=6.37×10^-8^), “positive regulation of immune system process” (UKB p=6.89×10^-192^, AoU p=9.48×10^-8^), and “positive regulation of cell activation” (UKB p=1.10×10^-80^, AoU p=4.3610^-7^). SCZ PRS association with CLBP/SDD case definitions was enriched for “positive regulation of signaling” in both UKB (p= 4.04×10^-7^) and AoU (p=3.45E×10^-6^). Top GO enrichments for other PRS associations included “bile acid secretion” (UKB-anxiety p=1.61×10^-21^), “cellular response to stress” (UKB-AUD p=7.67×10^-46^), “response to L-leucine” (UKB-PTSD p=1.25×10^-7^).

To translate GO enrichments into possible molecular targets, we conducted a drug repurposing analysis. After Bonferroni multiple testing correction (p<3.82×10^-5^), we identified several candidate drugs (Table S12). For depression PRS, we observed Bonferroni significant results in UKB and AoU. Although none of them survived multiple testing correction in both cohorts, we observed Bonferroni significant result in one cohort and nominal replication in the other for H-7 (UKB-depression p=1.05×10^-7^, AoU-depression p=0.023), fisetin (UKB-depression p= 2.25×10^-6^, AoU-depression p=0.002), mepenzolate bromide (UKB-depression p= 9.47×10^-6^, AoU-depression p=0.023), cephaeline (UKB-depression p= 1.15×10^-5^, AoU-depression p=0.011), HC toxin (AoU-depression p=2.47×10^-5^, UKB-depression p=0.007), and vorinostat (AoU-depression p=3.33×10^-5^, UKB-depression p=0.021). The latter compound was also Bonferroni-significant enriched in SCZ PRS (UKB p=1.63×10^-5^). Clozapine was also enriched for SCZ PRS (UKB p=8.94E-06). AUD PRS associations with CLBP/SDD case definitions were linked to naftidrofuryl (UKB p=2.08×10^-6^), perphenazine (UKB p=9.90×10^-6^), and proadifen (UKB p=1.62×10^-5^).

## DISCUSSION

The present study investigated 559,487 participants to assess the extent to which psychiatric disorders and their genetic predisposition contribute to CLBP-SDD comorbidity patterns. In line with previous evidence,^9, 10, 44–46^ we observed that psychiatric disorders are mostly associated with increased CLBP-SDD. However, integrating observational and genome-wide data, we uncovered that psychiatric disorders are linked to different CLBP-SDD comorbidity patterns. Our findings highlighted that these relationships could be explained by both direct effects and shared molecular pathways linking psychiatric disorders to CLBP-SDD comorbidity patterns.

Among the mental illnesses investigated, internalizing disorders (anxiety, depression, and PTSD) showed the strongest associations with CLBP and SDD in both UKB and AoU cohorts. In the observational analyses in UKB, anxiety and depression were more strongly associated with CLBP+SDD, while PTSD was more strongly associated with CLBP-only than CLBP+SDD. AoU observational analysis confirmed the stronger association of anxiety and depression with CLBP+SDD, but highlighted a different pattern for PTSD. PTSD was associated with increased CLBP+SDD and decreased SDD-only with the relationship with CLBP-only not surviving multiple testing correction. PTSD difference between UKB and AoU results may be due to how this disorder is differentially recorded in these healthcare systems. For instance, previous UK survey statistics highlighted potential barriers with one in eight of those who screened positive for PTSD having been diagnosed by a health professional.^47^ In with the possible issues related to PTSD phenotyping, our PRS analysis (which is based on genetic information rather than phenotype definitions) showed the same association pattern of internalizing PRS (including PTSD PRS) in both UKB and AoU with the strongest effect observed for CLBP+SDD. Interestingly, the multivariable PRS analysis indicated that genetic liability to depression was more strongly associated with CLBP/SDD case definitions (with the largest effect with respect to CLBP+SDD) than other psychiatric PRS. To understand whether these associations were due to causal effects or shared pathogenesis, we conducted further analyses. In UKB, our one-sample MR confirmed that the genetic liabilities to anxiety, depression, and PTSD are associated with increased CLBP/SDD case definitions. However, the strongest effects were observed with respect to SDD-only. In AoU, the effect sizes were much smaller than the ones observed in UKB, and they showed a slightly stronger effect with respect to CLBP+SDD. Although the sample size difference between UKB and AoU (N=325,975 vs. 48,994, respectively) may have contributed to distinct patterns observed, cohort-specific dynamics may also play a role in the results of the one-sample MR analysis. In the context of previous literature that highlighted the impact of internalizing disorders on pain sensitivity and chronic back pain,^48–54^ our study demonstrated that anxiety, depression, and PTSD and their genetic risk are associated with different CLBP/SDD comorbidity patterns, also highlighting that these relationships can vary across population groups and healthcare systems.

Comparing the effect sizes observed in observational, PRS, and one-sample MR analyses, it appears that direct effects of internalizing disorders and their genetic risk cannot fully explain their associations with CLBP/SDD case definitions. Our pathway-based analyses highlighted several biological processes, molecular functions, and cellular components shared between psychiatric disorders and CLBP/SDD case definitions. For depression, we identified five GO terms that reached Bonferroni correction in both UKB and AoU. Three of them (i.e., “positive regulation of immune system process”, “chemical homeostasis”, and “positive regulation of cell activation”) were related to immune systems and regulatory processes. These results align with transcriptomic and observational evidence suggesting that modulation of immune responses influences the development of chronic back pain.^55^ In line with previous hypotheses,^56–58^ these findings also support that immune pathways and processes regulating the homeostasis of biological systems are mechanisms linking depression and pain-related outcomes. The other two Bonferroni-significant GO terms were related to developmental processes (“developmental growth” and “generation of neurons”). Neurodevelopmental processes have been proposed in the pathogenesis of depression.^59^ Similarly, altered developmental processes have been highlighted as key contributors to spine-related disorders and back pain.^19, 60–62^ Our pathway-based analyses support that genes involved in developmental processes can act on different biological systems to contribute to the predisposition to depression, CLBP, and SDD.

The drug-repurposing analysis identified compounds that target the shared pathways linking depression to CLBP and SDD. Among the ones replicated between the two cohorts investigated, fisetin (a naturally occurring flavonoid) has shown evidence of neuroprotective, antidepressant, anti-inflammatory, and analgesic properties^63–66^ together with possible functional recovery after spinal cord injury.^67^ Two of the replicated compounds are histone deacetylase (HDAC) inhibitors (i.e., vorinostat and HC toxin). This may be related to the potential protective effect of HDAC inhibitors on neuroplasticity impairments^68^ and neuropathic pain.^69, 70^ Additionally, antidepressant activity has been reported for vorinostat in a chronic stress mouse model.^71^

Beyond internalizing disorders, we also observed the effect of AUD and its genetic risk on increasing CLBP and SDD with differences across the three case definitions less evident than the ones described above. These findings are in line with previous studies reporting how alcohol misuse can increase back pain^45, 72^ and negative spine-related outcomes.^73, 74^ Importantly, we observed discordant cross-ancestry associations in UKB observational analysis. Specifically, while the analysis in the whole cohort and most ancestry groups showed that AUD increases CLBP/SDD case definitions, AUD was associated with reduced CLBP-only and SDD-only in UKB participants of East Asian descent and reduced CLBP-only in UKB participants of Middle Eastern descent. To our knowledge, no information is currently available regarding the interplay among AUD, CLBP, and SDD in minority populations. Nevertheless, the inverse relationships observed may be partially due to health inequities and minority discrimination associated with AUD.^75^ Additionally, sociocultural factors (e.g., alcohol prohibition among Muslims) may also play a role. With respect to AUD, our genetically informed analyses highlighted that both direct effects and shared pathogenesis contribute to its association with CLBP/SDD case definitions. In particular, we uncovered those genes involved in “cellular response to stress” may be implicated in the pathogenesis of AUD, CLBP, and SDD, consistent with evidence linking cellular stress to these outcomes.^76–78^

In both UKB and AoU, we also observed a SCZ-specific pattern of associations. Specifically, SCZ was inversely related to CLBP/SDD case definitions. In the observational analysis, this inverse relationship was nominally significant but reached multiple testing correction in some ancestry groups (i.e., Central/South Asian and Admixed-Americans; Table S5). In the PRS analysis conducted only in EUR participants, SCZ genetic liability was inversely associated with CLBP/SDD case definitions in both UKB and AoU, especially when accounting for polygenic risk of other psychiatric disorders. These findings are in line with previous studies reporting that SCZ patients have increased pain tolerance, sensory threshold, and pain threshold, as well as reduced physiological response to noxious stimuli.^79^ Additionally, SCZ patients perceive nociceptive stimuli as less intense compared to healthy controls.^79, 80^ This altered pain response may lead to a lower likelihood of seeking medical attention for conditions typically associated with CLBP and SDD. Another possible dynamic contributing to our finding is that SCZ patients may express pain differently because of deficits in social skills, language, and communication.^14^ These may result in them being less likely to report accurate information to receive treatment for CLBP and SDD. SCZ drug repurposing analysis converged on vorinostat, which we also identified with respect to depression. Previous studies highlighted how HDAC inhibitors can reduce SCZ-like phenotypes and reduce side effects of antipsychotic medications.^81, 82^

While our findings expand the understanding of the impact of mental health on CLBP and SDD, we must acknowledge three limitations. First, the relationship of CLBP and SDD with psychiatric disorders is likely bidirectional as suggested in several studies.^9, 44^ In the present study we could not investigate CLBP and SDD effects on mental health using genetically informed approaches because of the lack of large-scale CLBP and SDD GWAS independent of the cohorts investigated (i.e., UKB and AoU). Second, our observational analyses highlighted possible differences in the interplay among psychiatric disorders, CLBP, and SDD in minority populations, but we could not follow up on these findings in our genetic analyses because of the lack of GWAS representative of diversity among human populations. Lastly, the two cohorts we included were not randomly sampled during participant enrollment, which may limit the representativeness. As a result, the generalizability of our findings to the broader global population may be limited.

In conclusion, the present study opens new directions in CLBP and SDD research, emphasizing the importance of investigating them in the context of mental health. Additionally, because of the direct effect and shared pathogenesis observed, our findings indicate that it may be clinically useful to screen for psychiatric comorbidities to understand the dynamics underlying CLBP/SDD comorbidity patterns. Importantly, we observed that minorities and sociocultural context may differentiate CLBP/SDD-related outcomes. Further studies are needed to follow up on this initial evidence. Similarly, our drug-repurposing analyses highlighted potential molecular targets that may be useful to identify therapeutics that simultaneously aim at the biological systems underlying the pathogenesis shared among psychiatric disorders, CLBP, and SDD.

## Data Availability

All data generated in the present study are reported in the manuscript and its supplemental material.

## Acknowledgments

The authors thank the participants and the investigators involved in the UK Biobank and the All of Us Research Program. This research has been conducted using the UK Biobank Resource (application reference no. 58146). The authors acknowledge support from the National Institutes of Health (RF1MH132337 and R33DA047527) and One Mind. The All of Us Research Program is supported by the National Institutes of Health, Office of the Director: Regional Medical Centers: 1 OT2 OD026549; 1 OT2 OD026554; 1 OT2 OD026557; 1 OT2 OD026556; 1 OT2 OD026550; 1 OT2 OD 026552; 1 OT2 OD026553; 1 OT2 OD026548; 1 OT2 OD026551; 1 OT2 OD026555; IAA #: AOD 16037; Federally Qualified Health Centers: HHSN 263201600085U; Data and Research Center: 5 U2C OD023196; Biobank: 1 U24 OD023121; The Participant Center: U24 OD023176; Participant Technology Systems Center: 1 U24 OD023163; Communications and Engagement: 3 OT2 OD023205; 3 OT2 OD023206; and Community Partners: 1 OT2 OD025277; 3 OT2 OD025315; 1 OT2 OD025337; 1 OT2 OD025276. In addition, the All of Us Research Program would not be possible without the partnership of its participants.

## Author Contributions

DQ, EF, MG, and RP designed the study. DQ and EF analyzed the data. JH, BCM, and MA supported the observational and genetic analyses. MA supported the epidemiological analyses. DQ and RP wrote the manuscript. All the other authors provided critical feedback, context interpretation, draft revision, and editing. RP supervised the study and received the primary funding that supported the study.

## Data sharing

All data generated in the present study are reported in the manuscript and its supplemental material. UKB data can be requested at https://www.ukbiobank.ac.uk/enable-your-research/apply-for-access. All of Us Research Program data are available at https://allofus.nih.gov/get-involved/opportunities-researchers.

