## Supplementary figures and images for "Understanding the comorbidities among psychiatric disorders, chronic low-back pain, and spinal degenerative disease using observational and genetically informed analyses"

### Supplementary Figure 1

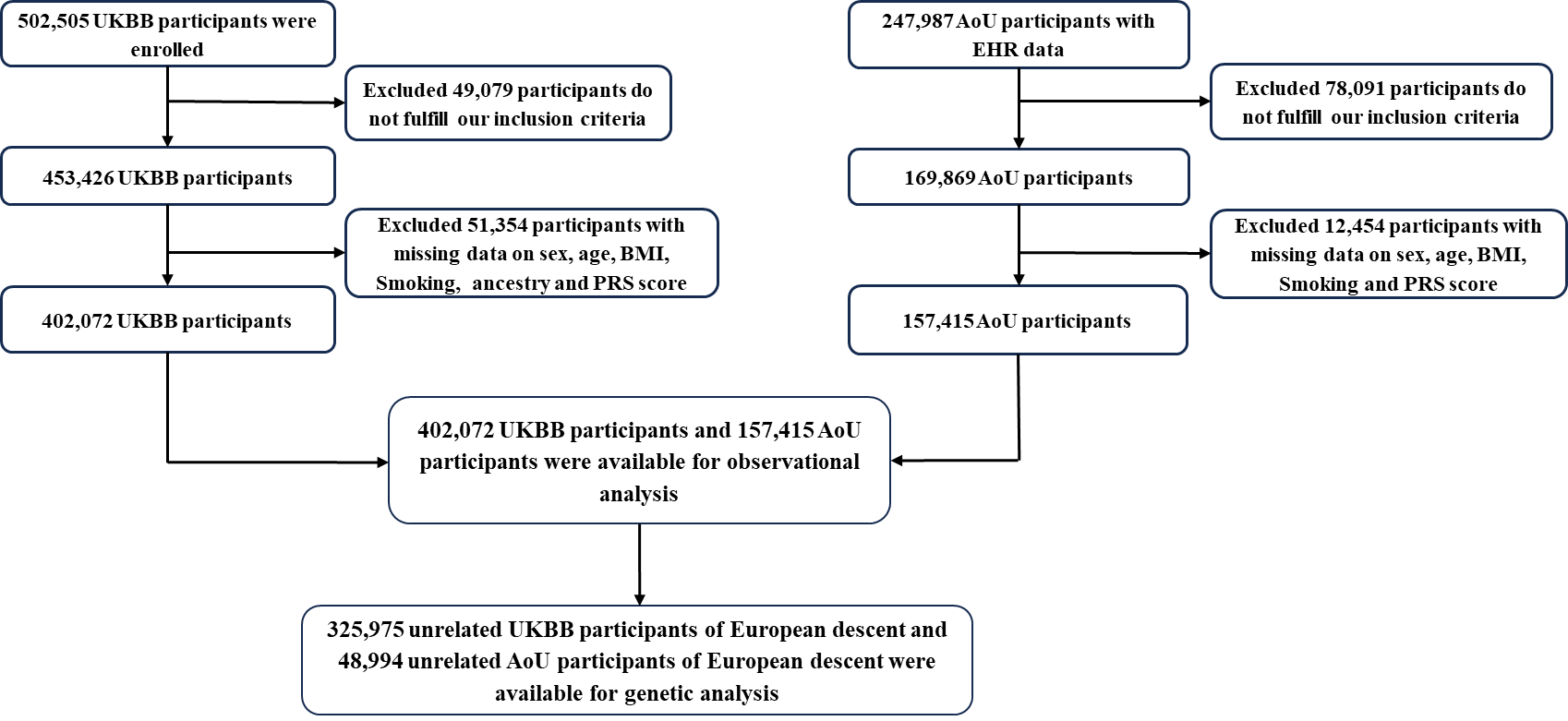

**Fig. S1 Sample size available from UK Biobank and All of US Research Program.**
